# Serial interval of COVID-19 and the effect of Variant B.1.1.7: analyses from a prospective community cohort study (Virus Watch)

**DOI:** 10.1101/2021.05.17.21257223

**Authors:** Cyril Geismar, Ellen Fragaszy, Vincent Nguyen, Wing Lam Erica Fong, Madhumita Shrotri, Sarah Beale, Alison Rogers, Vasileios Lampos, Thomas Byrne, Jana Kovar, Annalan M D Navaratnam, Parth Patel, Robert W Aldridge, Andrew Hayward, on behalf of Virus Watch Collaborative

## Abstract

**Introduction:** Increased transmissibility of B.1.17 variant of concern (VOC) in the UK may explain its rapid emergence and global spread. We analysed data from putative household infector - infectee pairs in the Virus Watch Community cohort study to assess the serial interval of COVID-19 and whether this was affected by emergence of the B.1.17 variant.

**Methods:** The Virus Watch study is an online, prospective, community cohort study following up entire households in England and Wales during the COVID-19 pandemic. Putative household infector-infectee pairs were identified where more than one person in the household had a positive swab matched to an illness episode. Data on whether individual infections were caused by the B.1.1.7 variant were not available. We therefore developed a classification system based on the percentage of cases estimated to be due to B.1.17 in national surveillance data for different English regions and study weeks.

**Results:** Out of 24,887 illnesses reported, 915 tested positive for SARS-CoV-2 and 186 likely infector-infectee pairs in 186 households amongst 372 individuals were identified. The mean COVID-19 serial interval was 3.18 (95%CI: 2.55 - 3.81) days. There was no significant difference (p=0.267) between the mean serial interval for Variants of Concern (VOC) hotspots (mean = 3.64 days, (95%CI: 2.55 – 4.73)) days and non-VOC hotspots, (mean = 2.72 days, (95%CI: 1.48 – 3.96)).

**Conclusions:** Our estimates of the average serial interval of COVID-19 are broadly similar to estimates from previous studies and we find no evidence that B.1.1.7 is associated with a change in serial intervals. Alternative explanations such as increased viral load, longer period of viral shedding or improved receptor binding may instead explain the increased transmissibility and rapid spread and should undergo further investigation.

## Introduction

The serial interval is defined as “the period of time between analogous phases of an infectious illness in successive cases of a chain of infection that is spread person to person” (Feinleib et al., 2001). Serial interval is often measured as the duration between symptom onset of a primary case and symptom onset of its secondary cases. This is a key epidemiological measure because it can allow investigation of epidemiological links between cases and it is an important parameter in infection transmission models used to inform infection control strategies. The doubling time of epidemic infections is in part dependent on both the serial interval and the R number (the average number of secondary infections each infection produces). Diseases with shorter serial intervals but similar R values will have shorter doubling times. Mean serial intervals vary widely for different respiratory infections and have been estimated at 2.2. days for influenza A H3N2, 2.8 days for pandemic influenza A(H1N1) pdm09, 7.5 days for respiratory syncytial virus, 11.7 days for measles, 14 days for varicella, 17.7 days smallpox, 18.0 days for mumps, 18.3 days for rubella and 22.8 days for pertussis (Vink et al., 2014).

Published estimates of the serial interval of COVID-19 are largely from Asian countries prior to the emergence of Variants of Concern (VOC). A meta-analysis of serial interval estimates for Covid-19 found mean serial intervals ranged from 4.2 to 7.5 days with a pooled mean of 5.2 (95%CI: 4.9–5.5) (Alene et al., 2021). A more recent concerning feature of COVID-19 epidemiology has been the emergence of a range of SARS-CoV-2 variants with mutations that may increase transmissibility, reduce the protective effective effect of immunity acquired from natural infection or vaccination and or increase clinical severity (Alene et al., 2021). These include B.1.1.7 (first described in England), 501Y.V2 (first described in South Africa) and, and P.1 (B.1.1.28.1 - first described in Brazil). Each of these variants rapidly became dominant in the country in which they were first described. For the B.1.17 variant increased transmissibility is thought to explain the rapid emergence and global spread. Since either increased R or decreased serial interval could potentially explain more rapid emergence of B.1.1.7, it is important to understand whether serial interval differs. To date, however there are no published comparisons of the serial interval for B.1.17 and previously circulating strains. We analysed data from putative household infector - infectee pairs in the Virus Watch Community cohort study to assess the serial interval of COVID-19 and whether this was affected by emergence of the B.1.17 variant.

## Methods

The Virus Watch study is an online, prospective, community cohort study following up entire households in England and Wales during the COVID-19 pandemic (Hayward et al., 2020). As of 11th April 2021, 49,149 people across England and Wales have joined the study. Participants prospectively complete detailed daily symptom diaries recording the presence and severity of any symptoms of acute respiratory, gastrointestinal and other illnesses. At the end of each week participants complete a weekly survey where they report any symptoms from that previous week as well as the dates and outcomes of any COVID-19 swabbing conducted as part of NHS Test and Trace, work-based testing schemes, and other research studies.

Symptom data were grouped into illness episodes. The start date of an illness episode was defined as the first day any symptoms were reported, and the end date was the final day of reported symptoms. A 7-day washout period where no symptoms were reported was used to define the end of one illness episode and the start of a new illness episode. Swab results were matched to illnesses that were within 14 days of each other. Putative household infector-infectee pairs were identified where more than one person in the household had a positive swab matched to an illness episode. Although negative serial intervals are possible, in practice it is not possible to assess the direction of transmission between pairs, so it was assumed that the minimum interval was zero days. According to clinical reports, the estimated latest possible transmission occurs 9 days after the infector’s symptom onset & the incubation period for the infectee can be up to 14 days (McAloon et al., 2020). Thus, the longest time interval between an infector’s and an infectee’s onset of symptoms was considered at 23 days. Our analysis considered pairs of cases with symptom onset occurring between 0 and 23 days apart in households as possible transmission pairs. Where there were multiple potential infectors for the same infectee, these pairs were excluded from the analysis (see Appendix). Serial interval was calculated as the number of days between symptom onset of the pairs of cases.

Data on whether individual infections were caused by the B.1.1.7 variant were not available. We therefore developed a classification system based on the percentage of cases estimated to be due to B.1.17 in national surveillance data for different English regions and study weeks. These surveillance data utilise a proxy indicator of B.1.17 known as Spike-gene target failure (SGTF) which can be picked up on most PCR assays used in English community testing programmes. Infections in regions and weeks when >75% of strains were SGTF were classified as occurring B.1.1.7 “hotspots”. Infections in regions and weeks when <25% of strains were SGTF were classified as occurring in “non-hotspots”. Infections in regions and weeks when 25% to 75% of strains were SGTF were classified as “undetermined” since no significant threshold was reached. Mean serial interval and 95% confidence intervals were compared in hotspot and non-hotspot areas using Welch Two Sample t-tests.

## Results

Out of 24,887 illnesses reported by 14,986 individuals within 9,991 households between 22/06/2020 and 07/03/2021, 7,304 were tested for SARS-CoV-2. Amongst the swabbed illnesses, 915 tested positive for SARS-CoV-2. 287 possible infector-infectee pairs in 194 households amongst 424 individuals were identified. Non-unique infectees (infectees with multiple potential infectors) were then excluded as their infectors could not be determined, resulting in 186 likely pairs in 186 households amongst 372 individuals between 03/09/2020 and 22/02/2021. 43 ‘infector-infectee’ pairs were identified in non-hotspot areas between 03/09/2020 - 05/12/2020, 69 were in hotspot areas between 14/12/2020 - 22/02/2021 and 74 in areas not determined between 09/09/2020 - 25/01/2021 (Table1/Table2).

**Table 1.** Demographic characteristics of the individuals included in the analysis.

| **VOC Status**<br>(Number of individuals) | **Overall**<br>(n = 372) |  | **Hotspot**<br>(n = 143) |  | **Non-hotspot**<br>(n = 85) |  | **Not determined**<br>(n = 144) |  |
| --- | --- | --- | --- | --- | --- | --- | --- | --- |
|  | n | % | n | % | n | % | n | % |
| **Sex (at birth)** |  |  |  |  |  |  |  |  |
| Female | 179 | 48.1 | 71 | 49.7 | 41 | 48.2 | 67 | 46.5 |
| Male | 170 | 45.6 | 68 | 47.6 | 42 | 49.4 | 60 | 41.7 |
| Missing | 23 | 6.3 | 4 | 2.7 | 2 | 2.4 | 17 | 11.8 |
| **Age (years)** |  |  |  |  |  |  |  |  |
| 0-15 | 25 | 6.7 | 11 | 7.7 | 5 | 5.9 | 9 | 6.2 |
| 16-44 | 114 | 30.6 | 45 | 31.5 | 26 | 30.6 | 43 | 29.9 |
| 45-64 | 165 | 44.4 | 63 | 44.0 | 29 | 34.1 | 73 | 50.7 |
| 65 + | 68 | 18.3 | 24 | 16.8 | 25 | 29.4 | 19 | 13.2 |
| **Region** |  |  |  |  |  |  |  |  |
| East Midlands | 46 | 12.4 | 8 | 5.6 | 20 | 23.5 | 18 | 12.5 |
| East of England | 56 | 15.1 | 36 | 25.2 | 2 | 2.3 | 18 | 12.5 |
| London | 62 | 16.7 | 44 | 30.7 | 9 | 10.6 | 9 | 6.2 |
| North East | 20 | 5.4 | 6 | 4.2 | 6 | 7.1 | 8 | 5.6 |
| North West | 54 | 14.5 | 11 | 7.7 | 16 | 18.8 | 27 | 18.7 |
| South East | 40 | 10.7 | 28 | 19.6 | 6 | 7.1 | 6 | 4.2 |
| South West | 16 | 4.3 | 2 | 1.4 | 6 | 7.1 | 8 | 5.6 |
| Wales | 10 | 2.7 | 0 | 0.0 | 0 | 0.0 | 10 | 6.9 |
| West Midlands | 18 | 4.8 | 6 | 4.2 | 4 | 4.7 | 8 | 5.6 |
| Yorkshire and The Humber | 28 | 7.5 | 2 | 1.4 | 16 | 18.8 | 10 | 6.9 |
| Missing | 22 | 5.9 | 0 | 0.0 | 0 | 0.0 | 22 | 15.3 |
| **Number of household members** |  |  |  |  |  |  |  |  |
| 2 | 214 | 57.5 | 71 | 49.6 | 55 | 64.7 | 88 | 61.1 |
| 3 | 84 | 22.6 | 41 | 28.7 | 12 | 14.1 | 31 | 21.5 |
| 4 | 54 | 14.5 | 25 | 17.5 | 12 | 14.1 | 17 | 11.8 |
| 5 | 16 | 4.3 | 2 | 1.4 | 6 | 7.1 | 8 | 5.6 |
| 6 | 4 | 1.1 | 4 | 2.8 | 0 | 0.0 | 0 | 0.0 |

**Table 2.**
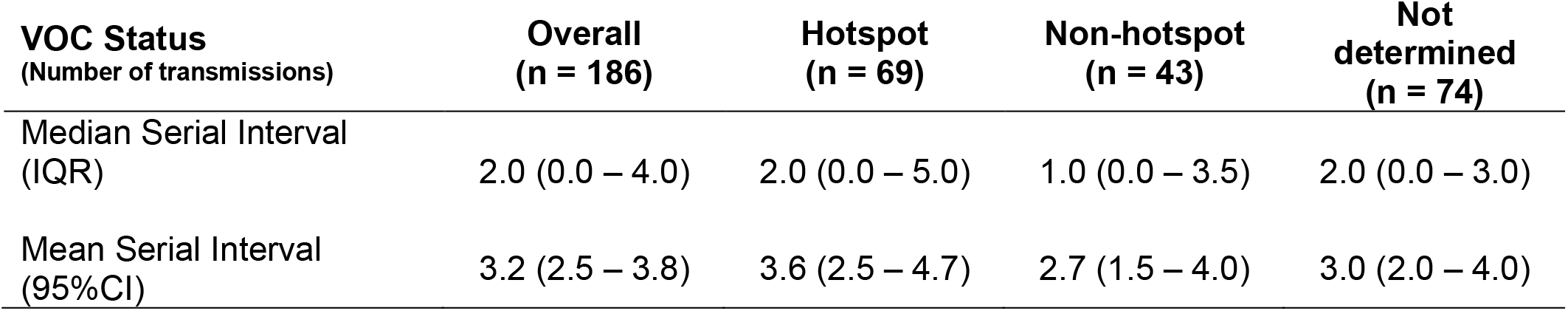
Serial Interval by VOC status.

| VOC Status<br>(Number of transmissions) | Overall<br>(n = 186) | Hotspot<br>(n = 69) | Non-hotspot<br>(n = 43) | Not<br>determined<br>(n = 74) |
| --- | --- | --- | --- | --- |
| Median Serial Interval<br>(IQR) | 2.0 (0.0 – 4.0) | 2.0 (0.0 – 5.0) | 1.0 (0.0 – 3.5) | 2.0 (0.0 – 3.0) |
| Mean Serial Interval<br>(95%CI) | 3.2 (2.5 – 3.8) | 3.6 (2.5 – 4.7) | 2.7 (1.5 – 4.0) | 3.0 (2.0 – 4.0) |

Figure 1 shows the distribution of serial intervals. The distribution peaks at 0.5 day and 90% of all observations lie between 0 and 8.5 days with values spanning up to 21 days. The mean COVID-19 serial interval was 3.18 (95%CI: 2.55 - 3.81) days and its median was 2 days (Table2).

**Figure 1.**
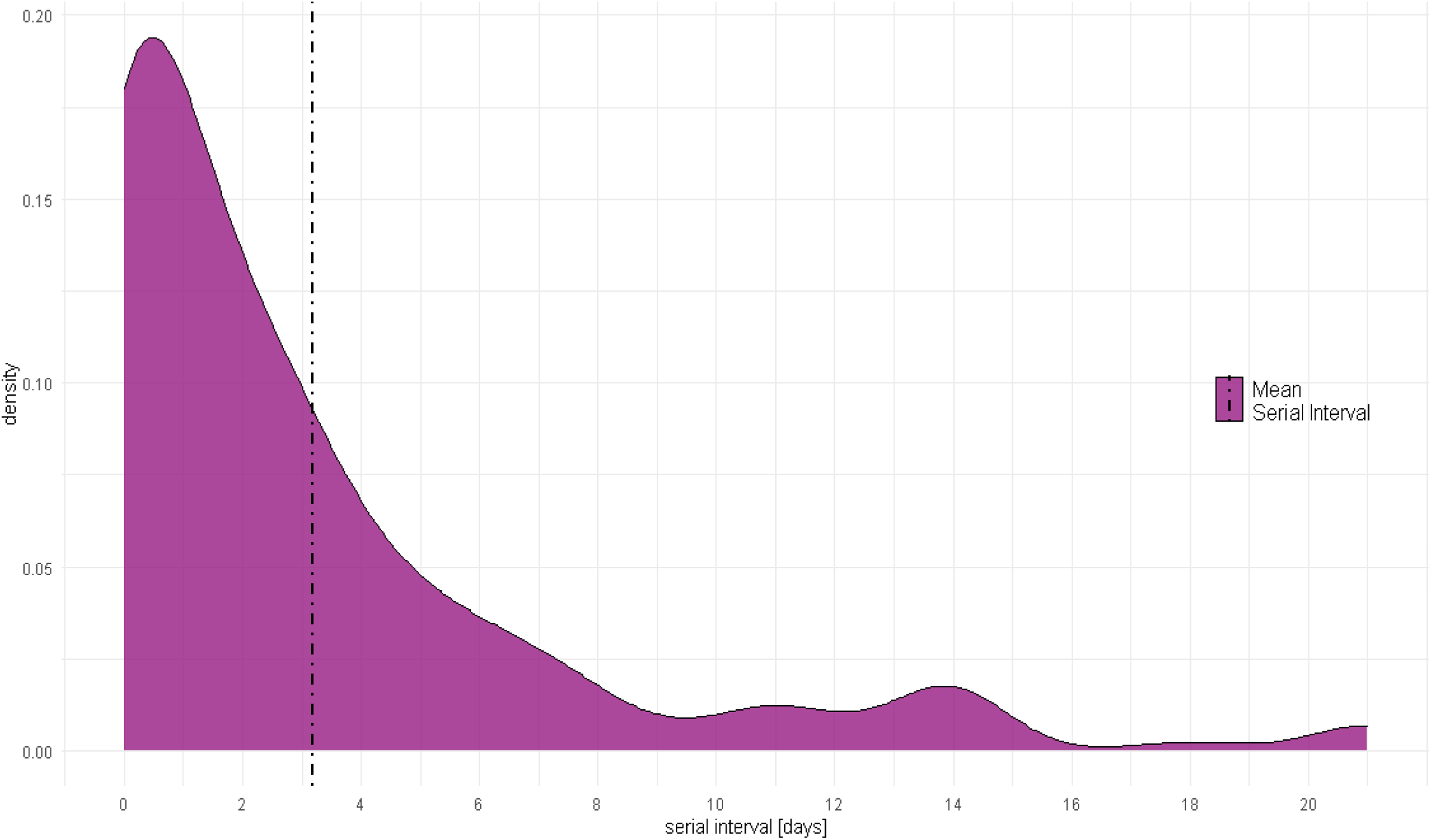
Serial Interval distribution and mean.

Figure 2 compares the distribution of serial intervals for B.1.1.7 hotspot areas and non-hotspot areas. There was no significant difference (p=0.267) between the mean serial interval for VOC hotspots (mean = 3.64 days (95%CI: 2.55 – 4.73), median = 2 days) and non-VOC hotspots (mean = 2.72 days (95%CI: 1.48 – 3.96), median = 1 day) (Table2).

**Figure 2.**
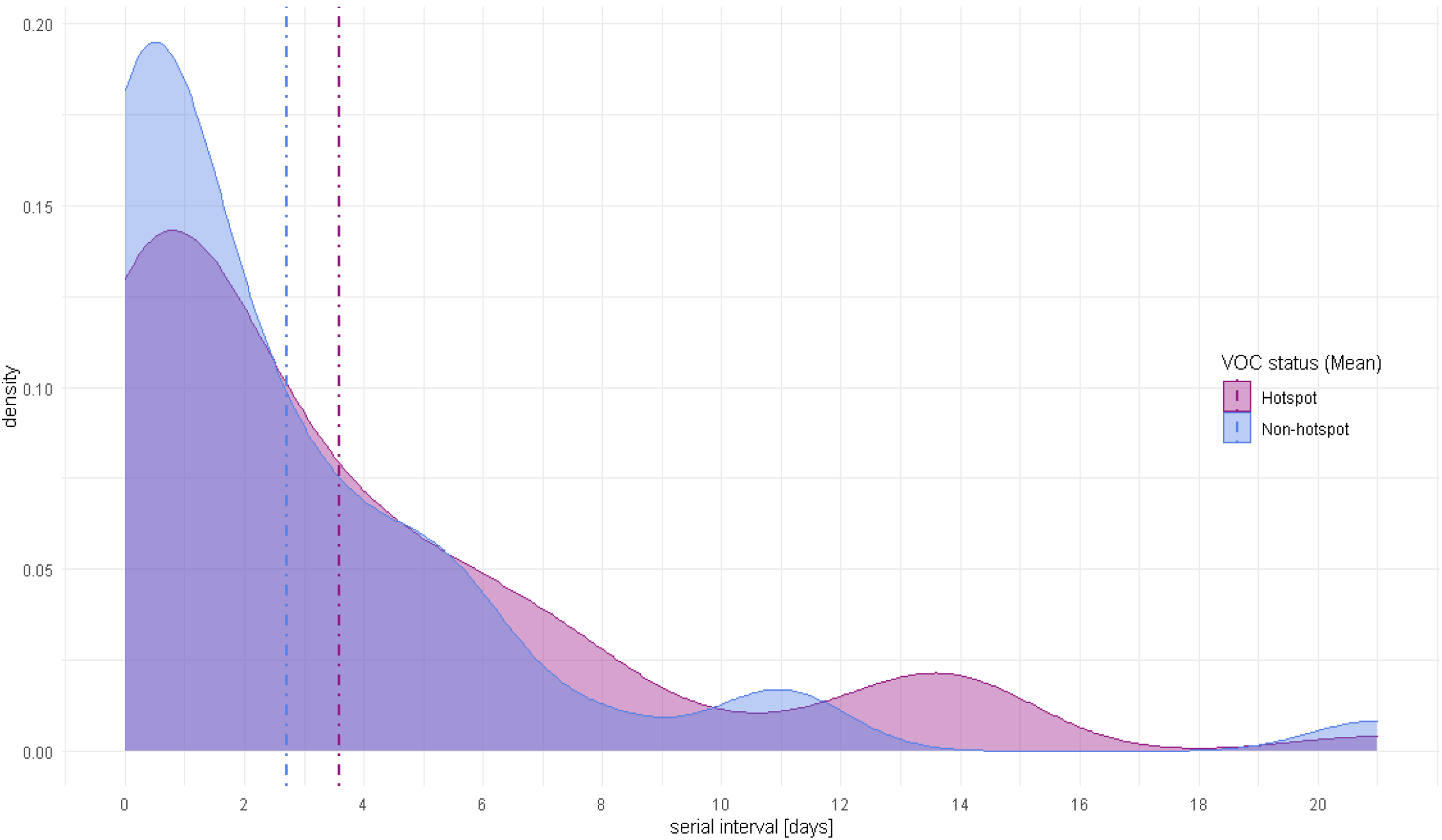
Serial interval density distribution for B.1.1.7 hotspot areas and non-hotspot areas (Mean).

## Discussion

Our estimate of the mean serial interval of COVID-19 (3.18 days - 95% CI 2.55 - 3.81) is within the range of previous studies reviewed by Griffin et al (2020), but slightly lower than pooled estimates from meta-analysis of data from international studies in the first few months of the pandemic (5.2 (95%CI: 4.9–5.5) (Alene et al., 2021). Differences in populations, social contact, and timeframes may explain the range of estimates reported. The implementation of control measures, regular testing, isolation and improved knowledge of SARS-CoV-2’s transmission since the start of the pandemic, may have reduced the potential for an infected person to transmit the disease over a long period of time. Multiple studies observed and attributed the decrease of the serial interval to increased control measures (Zhao et al.,2020; Bi et al., 2020; Lavezzo et al., 2020). Ali et. al (2020) modelled the serial interval over time accounting for timeliness of cases’ isolation and found that “serial intervals are positively associated with isolation delay”. Another potential explanation of shorter serial intervals may be due the frequent and close contact among household members. This could lead to transmissions occurring earlier in the course of infection which would result in shorter serial intervals.

Strengths of the study include the relatively large number of pairs compared to most studies, the prospective daily recording of symptoms and weekly reporting of swab test results in a large household cohort, and our ability to assess whether a variant with apparent increases in transmissibility has an altered serial interval. Limitations of our analysis include reliance on samples taken during the national symptomatic testing programme to assess infection meaning we are likely to have missed some infections and household transmission events. Pooled asymptomatic proportion of SARS-CoV-2 infections is estimated at 23% (95% CI 16%-30%) and we cannot assess serial intervals when either case is asymptomatic (Beale et al., 2020). We can also not assess the possibility of negative serial intervals which may arise when transmission occurs prior to symptom onset and incubation period is short. Finally, we do not have individual information on whether strains were B.1.17 and used a proxy measure of this based on levels of SGTF in different areas and times. These limitations reduce the likelihood of observing differences between B.1.17 other wild-type strain types at the time of the study.

Our analysis does not provide evidence to suggest that changes in serial interval explain the rapid emergence of B.1.17. Alternative explanations such as increased viral load (Kidd et.al, 2021) or improved receptor binding may instead explain the increased transmissibility and rapid spread and should undergo further investigation.

## Data Availability

We aim to share aggregate data from this project on our website and via a "Findings so far" section on our website - https://ucl-virus-watch.net/. We will also be sharing individual record level data on a research data sharing service such as the Office of National Statistics Secure Research Service. In sharing the data, we will work within the principles set out in the UKRI Guidance on best practice in the management of research data. Access to use of the data whilst research is being conducted will be managed by the Chief Investigators (ACH and RWA) in accordance with the principles set out in the UKRI guidance on best practice in the management of research data. We will put analysis code on publicly available repositories to enable their reuse.

## Ethics

This study has been approved by the Hampstead NHS Health Research Authority Ethics Committee. Ethics approval number - 20/HRA/2320.

## Data availability

We aim to share aggregate data from this project on our website and via a “Findings so far” section on our website https://ucl-virus-watch.net/. We will also be sharing individual record level data on a research data sharing service such as the Office of National Statistics Secure Research Service. In sharing the data, we will work within the principles set out in the UKRI Guidance on best practice in the management of research data. Access to use of the data whilst research is being conducted will be managed by the Chief Investigators (ACH and RWA) in accordance with the principles set out in the UKRI guidance on best practice in the management of research data. We will put analysis code on publicly available repositories to enable their reuse.

## Appendix A: supplementary methods description

**Figure 3.**
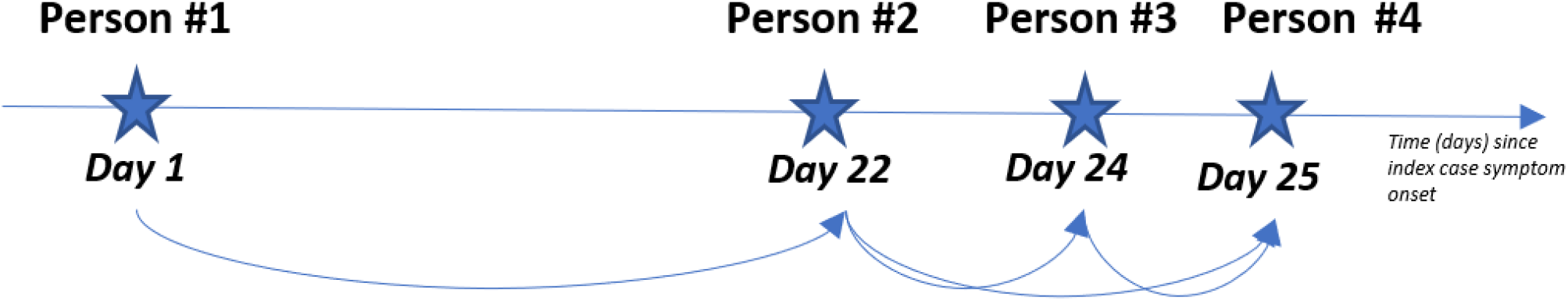
Household transmission dynamics diagram. Consider household X with 4 confirmed COVID-19 cases and their respective symptom onset date. Any case with a symptom onset date within 23 days of a previous case will be paired. Where there were multiple potential infectors for the same infectee, these pairs were excluded from the analysis ‘Person #4’ has two potential infectors as her symptom onset date is within 23 days of ‘Person #2’ and ‘Person #3’ ‘s symptom onset dates. Pairs containing ‘Person #4’ as an infectee will be removed since we cannot determine her “true” infector. Thus, for our analysis, we will only retain the two most likely transmission pairs: Person #1 -> Person #2 Person #2 -> Person #3

